# Multi-polygenic scores for externalizing behavior in schoolchildren

**DOI:** 10.64898/2025.12.19.25342558

**Authors:** Uxue Zubizarreta-Arruti, María Soler Artigas, Rosa Bosch, Judit Cabana-Domínguez, Natalia Llonga, Pau Carabí-Gassol, Valeria Macias-Chimborazo, Manuel D. Rodríguez Romero, Mireia Pagerols, Elia Pagespetit, Raquel Prat, Júlia Puigbo, Josep Antoni Ramos-Quiroga, Miquel Casas, Silvia Alemany, Marta Ribasés

## Abstract

**Background:** Externalizing behavior refers to emotional and behavioral problems or disorders characterized by conducts directed outward at an individual’s environment. Polygenic scores (PGSs) indexing the individual genetic susceptibility for this behavior still explain a small proportion of the phenotypic variance. To increase this phenotypic variance explained for externalizing behavior we used a multi-PGS approach combining PGSs for several risk factors, mental health conditions and related phenotypes. In addition, we assessed the potential moderating effect of socioeconomic status (SES).

**Methods:** The study included a total of 4,485 children and adolescents (mean age = 10.0 years; range = 5-18, 45% females) with behavioral and genetic data available. Externalizing behavior was assessed using the Child Behavior Checklist and PGSs were constructed using PRScs software. We tested two models and compared the proportion of variance that they explained: (i) a single-PGS model with the PGS for externalizing behavior (PGSexternalizing) and (ii) a multi-PGS model combining the PGSexternalizing with other PGSs for risk factors grouped in six categories: environment, mental-health, cognition, personality, health-risk behaviors and non-mental diseases.

**Results:** The multi-PGS models for the categories of environment, mental-health, cognition and health-risk behaviors improved the variance explained of externalizing behavior in 20.9%, 37.6%, 35% and 34.2%, respectively, over the single model only including PGSexternalizing. Furthermore, we observed significant interactions between SES and the multi-PGS models of environment, mental-health, cognition and health-risk behaviors (P-interaction < 0.05), indicating that in individuals with lower SES, the influence of these four multi-PGSs on externalizing behavior is stronger.

**Conclusion:** This study provides new insights into the genetic architecture of externalizing behavior in a population-based cohort of children and adolescents and further supports the utility of incorporating multiple PGSs into predictive models to improve accuracy, increase the proportion of variance explained, and maximize the predictive utility of PGSs. Additionally, by assessing GxE, we contribute to understanding the complex relationship between SES and mental health outcomes, highlighting its impact on youth.

## INTRODUCTION

Approximately 75% of mental disorders onset by childhood, adolescence or early adult life (Kessler et al., 2005). Most studies suggest that 10-20% of children and adolescents are affected by mental-health disorders annually (Ogundele, 2018). Externalizing behavior, in particular, refers to emotional and behavioral problems or disorders characterized by conducts directed outward at an individual’s environment and causes significant suffering, not only to the individuals affected but also to their families, communities, and society at large (Thapar & Riglin, 2020). It comprises a broad range of behaviors and/or disorders including substance use disorders (SUD), conduct disorder and attention deficit/hyperactivity disorder (ADHD), among others (Ogundele, 2018; Krueger et al., 2002; Young et al., 2000). These behaviors tend to manifest early in life, with an age of onset around 10 and 15 years for ADHD and SUD, respectively (McGrath et al., 2023; Barr & Dick, 2020).

Externalizing behavior is a complex trait with both genetic and environmental factors involved in its etiology (Faraone & Larsson, 2019; Polderman et al., 2015). Indeed, twin and family studies show a strong genetic influence underlying externalizing behavior, with reported heritability estimates around 80% (Hicks et al., 2004). This high heritability estimate makes it a good target for genome-wide association studies (GWASs), which allow to identify single nucleotide polymorphisms (SNPs) that increase the likelihood to develop a particular disorder or trait and can be used to construct genome-wide polygenic scores (PGSs), which index an individual’s overall genetic liability to develop the trait of interest. The largest GWAS on externalizing behavior was conducted in around 1.5 million individuals, identifying more than 500 risk loci (Karlsson Linnér et al., 2021). Also, PGS analyses showed evidence of association between the genetic liability to externalizing behavior and a wide range of traits and disorders (Abdellaoui & Verweij, 2021). For instance, Linner et al. reported association between PGS for externalizing behavior (PGS_externalizing_) and a broad variety of behavioral and social outcomes related to self-regulation, such as different lifetime substance use, psychiatric disorders or criminal justice system involvement (Karlsson Linnér et al., 2021).

Although PGSs have proven to be a useful tool for understanding genetic liability, even in large cohorts, they continue to explain only small portions of the variance of the trait of interest in independent samples (Derks et al., 2022). For externalizing behavior, PGS_externalizing_ explained 8.9 and 10.5% of its variance in two independent samples (Karlsson Linnér et al., 2021). As an alternative for single-trait PGS models, multi-polygenic score (multi-PGS) approaches have been described as a promising method to increase variance explained. The multi-PGS approach combines multiple single-trait PGSs using penalized regression models and selecting only PGSs for traits that improve prediction accuracy (Krapohl et al., 2018; Albiñana et al., 2023). Using this study design, Krapohl et al. improved the prediction accuracy for educational achievement, general cognitive ability and body mass index (Krapohl et al., 2018). Similarly, by using multi-PGSs, Albiñana et al. increased the prediction accuracy for ADHD and autism spectrum disorder up to 9-fold in terms of variance explained over single-PGS models (Albiñana et al., 2023).

Beyond genetic factors, environmental influences, such as socioeconomic status (SES), play a key role in mental health. Previous studies report that children from families with lower SES are more likely to exhibit higher levels of psychopathology, including externalizing symptoms (Bøe et al., 2014). Additionally, both low and decrease in SES are major predictors of the onset of mental-health problems in children and adolescents (Reiss, 2013). Some studies conducted on externalizing problems also examined gene-environment interaction (GxE). For instance, findings on two independent pediatric samples show that when the genetic load for schizophrenia, bipolar disorder, cannabis use disorder and alcohol dependency is low, ADHD and inattention symptoms are more strongly influenced by family income (Mooney et al., 2023). However, Mooney et al. reported that these effects were not replicated.

In the present study we aim to increase the proportion of variance for externalizing behavior explained by PGS_externalizing_ using a multi-PGS approach combining traits for multiple risk factors and mental-health phenotypes in a population-based cohort of children and adolescents. In addition, in order to better understand the role of SES in mental health, we tested gene-environment interactions by assessing the moderation effects of SES in the relationship between multi-PGSs and externalizing behavior.

## METHODS

### Sample

The present study included 4,485 participants aged 5 to 18 years and recruited between 2013 and 2019 from 48 schools in Catalunya, Spain. All participants were of European descent and collected from the population-based INSchool initiative (Bosch et al., 2022). All individuals had data available on genetics, socioeconomic status, demographic and behavioral outcomes assessed at time of recruitment. The study was granted a full ethics approval by the ethic committees of the Hospital Universitari Vall d’Hebron and the Hospital Materno-Infantil Sant Joan de Déu (Catalonia, Spain). Written consent was obtained from all parents/caregivers.

### Outcome assessment

Externalizing behavior was assessed using the Child Behavior Checklist (CBCL) (Achenbach, 1991; Achenbach, 2001). The CBCL has a total of 112 items about the child’s functioning during the past 6 months, which caregivers answered by using a 3-point Likert scale (0 = Not True, 1 = Somewhat or Sometimes True, 2 = Very True or Often True). These answers are then used to calculate summary scores that can be grouped into broadband and syndrome scales. In the current study, we examined the externalizing broadband scale, which combines items from the rule-breaking behavior and aggressive behavior syndrome scales. Higher scores indicate more behavioral problems.

### Socioeconomic status assessment

The Hollingshead Four-Factor Index (Hollingshead, 2011) was used to measure individuals’ SES, which was computed by weighting parents’ responses to a questionnaire about their education level and occupation. The resulting scores range from 8 to 66, with higher values indicating higher SES.

### Genotyping, imputation and quality control

Genomic DNA was isolated from saliva samples or buccal swabs collected using Oragene DNA OG-500 or OC-175 kits, respectively (DNA Genotek) and genotyped in three genotyping waves: Illumina Infinium PsychChip_v1.0 array for wave 1 (n = 810), the Infinium Global Screening Array-24 version_2 (GSA_v2) for wave 2 (n = 2,806) and the Infinium Global Screening Array-24 version_3 (GSA_v3) for wave 3 (n = 869) (Illumina, CA, San Diego, USA). Pre-imputation quality control was done with the PLINK 2.0 (Chang et al., 2015) and included individual and variant filtering based on the following parameters: variant call rate > 0.95 (before individual filtering), individual call rate > 0.98, autosomal heterozygosity deviation (|Fhet| < 0.2), variant call rate > 0.98 (after individual filtering), SNP Hardy-Weinberg equilibrium (P > 1 x 10^-6^) and minor allele frequency (MAF) > 0.01. Genetic outliers were identified by principal component analysis (PCA) using PLINK 2.0 and the mixed ancestry 1000G reference panel (Auton et al., 2015). Ancestry outliers were excluded if their principal component (PC) values for PC1 or PC2 were greater than 1 standard deviation from the mean-centering point for our sample, considering each GWAS wave separately. Imputation was done with McCarthy tools, for data preparation, and the Michigan Imputation Server, using the Haplotype Reference Consortium (HRC_Version_r1.1_2016) reference panel (GRCh37/hg19). Post-imputation quality control per wave included imputation INFO score ≥ 0.8 and MAF > 0.01 and variant call rate > 0.95. SNPs present in all GWAS waves were considered and related or duplicate individuals were removed with the “KINGrobust kinship estimator” analysis with PLINK 2.0 considering a kinship coefficient exceeding 0.0442. We applied a final individual call rate threshold of 0.95. We converted the dosage files into best guess files with PLINK 2.0 and filtered for MAF > 0.01 and call rate > 0.95 for downstream PGS analyses (n_SNP_ = 3,698,091).

### Polygenic Scores

Genome-wide polygenic scores were constructed using PRScs software (Ge et al., 2019) and PLINK 1.9. For the primary trait of study, externalizing behavior, PGS_externalizing_ was computed using summary statistics from the largest GWAS available to date (Karlsson Linnér et al., 2021). To construct hypothesis-driven multi-PGSs we searched for publicly available GWAS’ summary statistics for traits and disorders grouped in six categories: environment, mental-health, cognition, personality, health-risk behaviors and non-mental diseases. They all must fulfill the following inclusion criteria: (i) non sex-specific; (ii) European ancestry; (iii) sample size [N_effective_ = 4 x N_cases_ x N_controls_ / (N_cases_ + N_controls_)] ≥ 5,000 and (iv) heritability Z-score ≥ 4 (obtained using LD score regression). When more than one dataset was available for a single trait, the one with the largest sample size was selected. After applying these inclusion criteria, a total of 66 traits were considered in the study (**Supplementary Table 1**). The resulting datasets, depending on their available data, were filtered out for indels, duplicates, minor allele frequency (MAF) > 0.01, INFO > 0.8 and SNP_Neffective_ > 70%. All PGSs were standardized to a mean of 0 and a standard deviation of 1.

### Statistical analysis

To improve our comprehension of the subsequent models, we performed Pearson’s correlation tests between the single-trait PGS within each of the six categories.

### Single- and multi-PGS models

We compared two models for externalizing behavior, the single- and the multi-PGS models. The base model was the single-PGS model, where PGS_externalizing_ was the only independent variable. Then, we constructed a multi-PGS for each of the six categories of study (i.e.; multi-PGS_environment_, multi-PGS_mental-health_, multi-PGS_cognition_, multi-PGS_personality_, multi-PGS_health-risk_behaviors_ and multi-PGS_non-mental_diseases_). Each multi-PGS model consisted of the PGS_externalizing_ along with the PGSs of the other traits in the category. To this aim, per each category, we constructed an L1 penalized regression (lasso), with the cv.glmnet function from the glmnet R package (Friedman et al., 2025), including as independent variables PGS_externalizing_ together with PGSs of all the traits in each category. In all lasso models, the options were alpha = 1, family = “gaussian” and the default penalty.factor of 1 for all variables, except for PGS_externalizing_, so it was not regularized. We selected the regularization parameter, lambda, corresponding to the minimum mean squared error (“lambda.min”), determined through 10-fold cross-validation.

For both the single- and multi-PGS analyses, associations were tested using linear regression models. In all models, the dependent variable was the residuals from a linear mixed-effects model with square-root transformed externalizing behavior score, to approximate normal distribution. This model was adjusted by sex, age and the first 10 genetic principal components (PCs) and school as random effect to account for the multilevel nature of the data.

Both single- and multi-PGS models were tested using a repeated 5-fold cross-validation design. We randomly split the sample into five equal-sized subsets of 897 individuals. For each iteration, we used four subsets as the training data (for training and optimizing the models) and the remaining subset as the validation data (to estimate trained models’ performance). The cross-validation process was repeated five times, where each of the five subsets was used once as the validation data. In addition, to minimize variation across validation sets, we repeated the 5-fold cross-validation five times with random data set partitions in each. The final contribution of each predictor PGS was calculated as the average of the weights obtained for that PGS across the 5 fold-cross validation repeated 5 times (25 iterations in total). The standard deviation (SD) across all the 25 estimates was used as a measure of dispersion.

### Prediction accuracy

In each validation set, the prediction accuracy of the model was calculated as out-of-sample R^2^ (R^2^_OF_) following equation 1, where y_testing_ are the aforementioned residuals from the linear mixed-effect model of the validation data, y_predicted_ are the predicted values obtained from the respective single-PGS or multi-PGS model and y_training_ are the residuals of the training data.

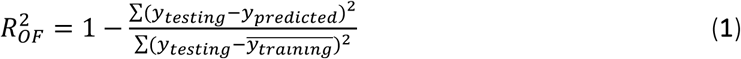

The relative improvement when comparing models was estimated as adjusted R^2^ (R^2^_adj_), defined as the variance explained by the multi-PGS model 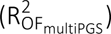 after accounting for the variance explained by the single-PGS model 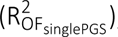, following equation 2.

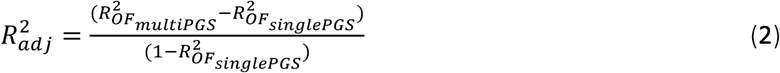

After completing all five repetitions, we calculated the average R²_adj_ per multi-PGS to obtain a single performance increase metric for each category. We also calculated the average improvement percentage per each multi-PGS considering the R^2^_OF_ of the single-PGS model 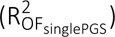 as reference.

Given that each category had a different number of traits and this may affect the proportion of variance explained, we used a permutation test to compare the R^2^_adj_ increase among categories. To this aim, we calculated the R^2^_adj_ for each multi-PGS model using permuted PGSs values. This process was repeated 100 times, generating an empirical null distribution of R^2^_adj_ assuming no effect of the PGSs on the phenotype. We then calculated the number of null distribution’s SD that the observed R^2^_adj_ was away from the mean of the null distribution.

### Moderating effects by SES

The main effect of SES on externalizing behavior was examined in the full sample using a linear model, with the residuals of externalizing behavior, from the linear mixed-effect model described above, as the dependent variable and SES as the independent variable.

We then assessed the potential moderating effect of SES on the association between externalizing behavior and the multi-PGSs showing improvement of the prediction accuracy of externalizing behavior. In these analyses, we also used a linear model with the residuals of externalizing behavior as the dependent variable and the main effects and interaction term for SES and the predicted values obtained in the multi-PGS models. For simplicity, the predicted values used in these models were the ones obtained in the first 5-fold cross-validation (first repetition) in the multi-PGS analyses.

For each category, we ran the model on each partition and results from the five partitions were pooled using fixed-effects inverse variance weighted meta-analyses using the meta R package (Schwarzer, 2025). To visualize results from the interaction analysis we plotted results from a single lineal regression model obtained considering the whole sample and adding the partition set as covariate.

## RESULTS

The mean age of the participants was 10 years (SD = 2.98) and approximately half of the sample were girls (45%) (**Table 1**). The average SES was 44.76 (SD = 12.57, min = 8, max = 63) and the mean score for externalizing behavior was 6.89 (SD = 6.35, min = 0, max = 62).

**Table 1.** Characteristics of the sample.

| **Characteristic** | **INSchool sample**<br>**(N = 4,485)** |
| --- | --- |
| Age [mean(min-max)] | 9.94 (5-18) |
| Sex (female), N (%) | 2,019 (45.02) |
| Socio-economic status [mean(min-max)] | 44.76 (8-63) |
| Externalizing behavior [mean(min-max)] | 6.89 (0-62) |

A total of 66 PGS within 6 categories were considered in the multi-PGS analysis. Some of them were highly correlated such as PGS for intelligence quotient and cognitive performance (r = 0.85, p-value < 2.2 x 10^-16^), and PGS for risk taking and risk tolerance (r = 0.93, p-value < 2.2 x 10^-16^) included in the multi-PGS_cognition_ and the multi-PGS_health-risk_behaviors,_ respectively (see correlation results in **Supplementary Figure 1**).

The PGS_externalizing_ showed positive association with externalizing behavior and yielded a mean weight of 0.12 (SD = 0.012), indicating that the higher the PGS_externalizing_, the higher score in externalizing behavior (**Figure 1, Supplementary Table 2**). When comparing the performance of the six multi-PGS models tested, four of them, multi-PGS_environment_, multi-PGS_mental-health_, multi-PGS_cognition_ and multi-PGS_health-risk_behaviors_, substantially improved the prediction accuracy of children’s externalizing behavior in 20.9%, 37.6%, 35% and 34.2%, respectively, over the single-PGS model only including PGS_externalizing_ (**Figure 2, Supplementary Table 3**).

**Figure 1.**
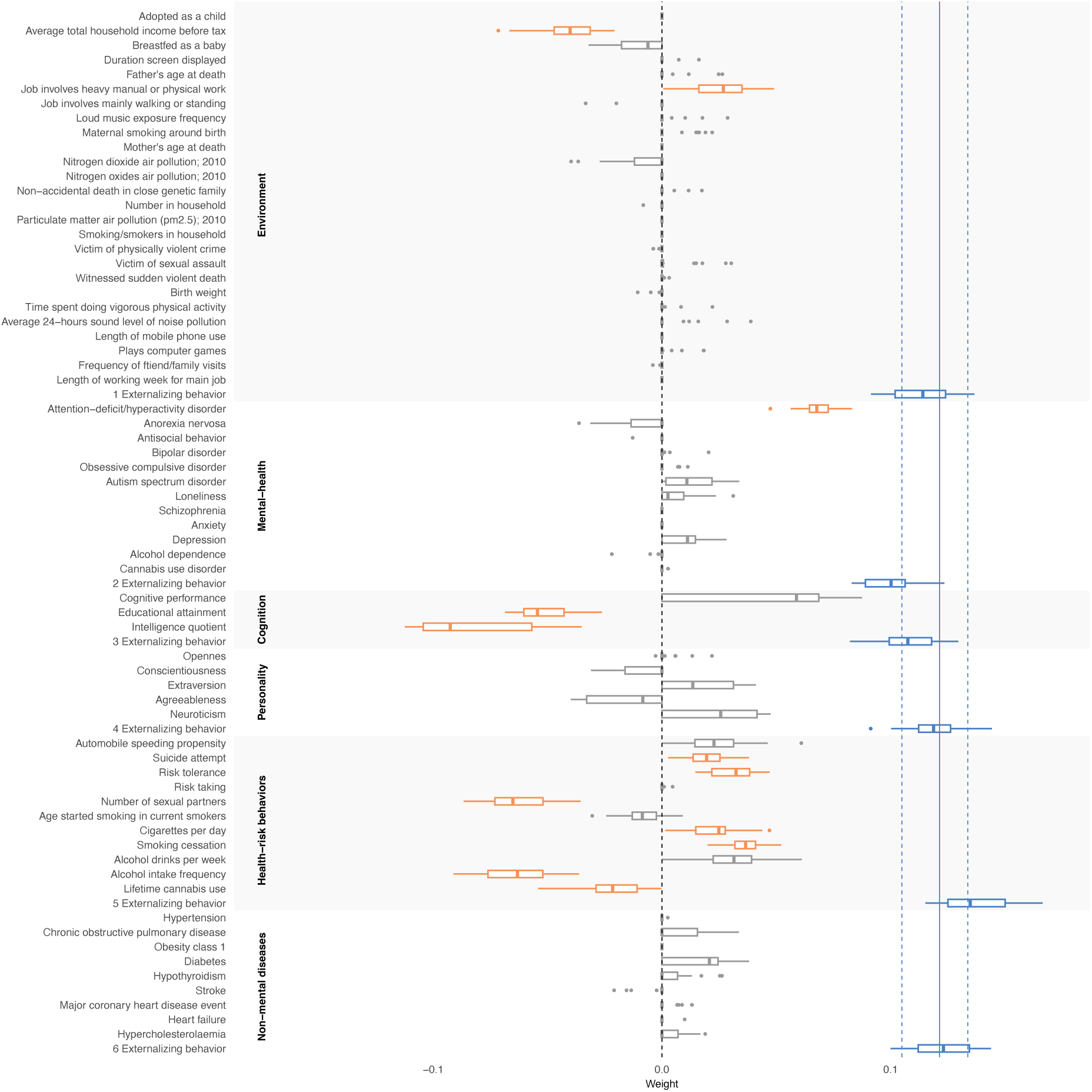
Boxplots for traits included in the environment, mental-health, cognition, personality, health-risk behaviors and non-mental diseases multi-PGS models. PGSs for traits contributing to increase the prediction accuracy of externalizing behavior with weights > 0 and consistent direction of effect across iterations are shown in orange and PGS_externalizing_ are presented in blue. The continuous blue solid line represents the mean weight and the dashed lines its maximum and minimum weight in the single-PGS model only including PGS_externalizing_.

**Figure 2.**
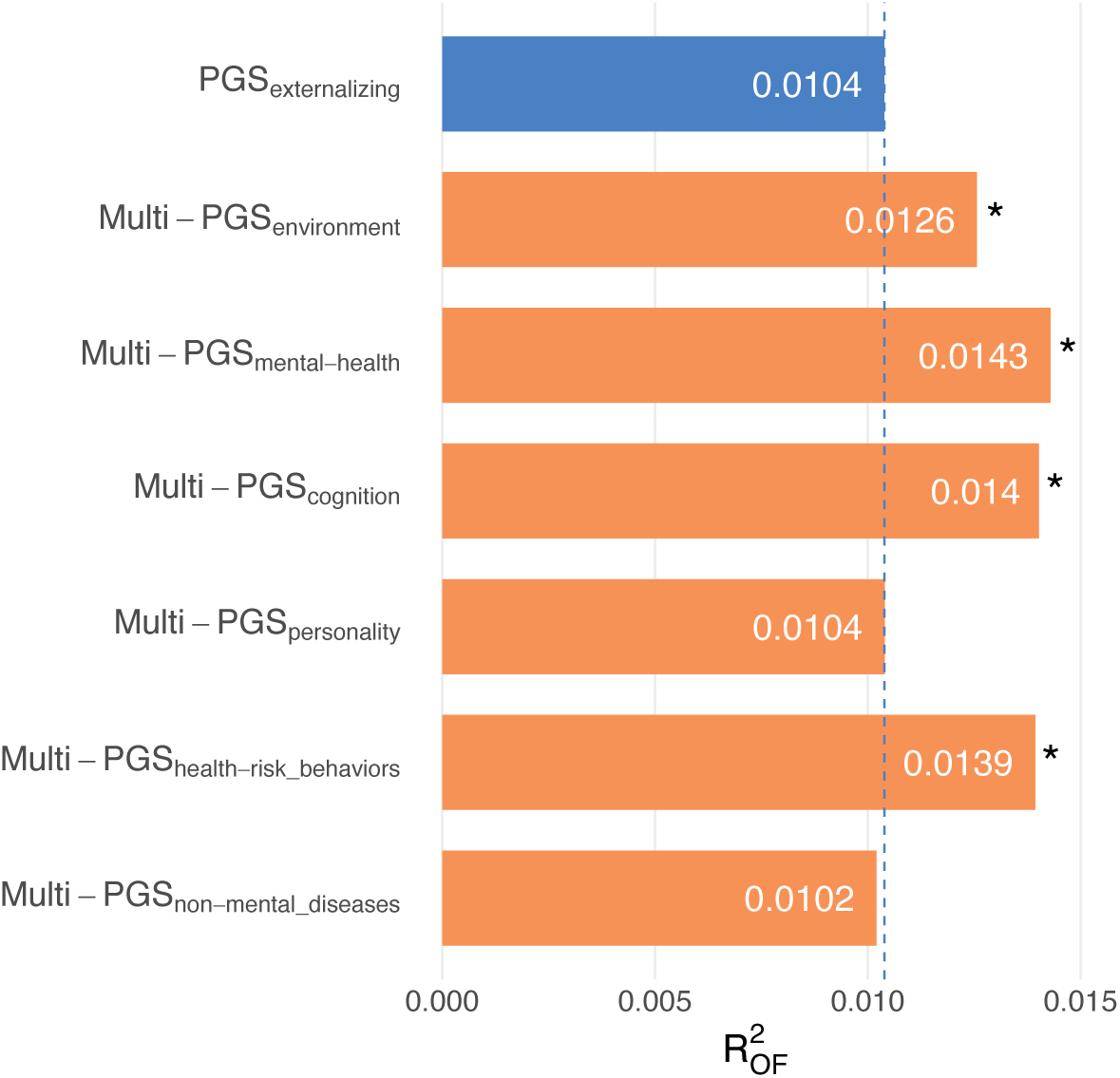
R^2^_OF_ of the single-PGS model including PGS_externalizing_ (in blue) and the six multi-PGS models by categories (in orange). Asterisks represent a distance of at least 9 SD between the observed R^2^_adj_ of the multi-PGS model and the mean R^2^_adj_ of the null distribution obtained in the permutation test assessed in the same multi-PGS model.

Although PGS_externalizing_ remained the most significant contributor to externalizing behavior across all the multi-PGS models tested (**Figure 1, Supplementary Table 2**), other PGSs also contributed consistently (always receiving a non-zero weight with a consistent sign) to the multi-PGS models and increased the prediction accuracy over the single-PGS model. For instance, the PGS for average total household income was negatively associated with externalizing behavior (β = −0.04; SD = 0.012), while PGS for job involves heavy manual or physical work was positively associated with externalizing behavior (β = 0.02; SD = 0.013), for the multi-PGS_environment_ model. PGS for ADHD was the strongest contributor for multi-PGS_mental-health_, also positively associated with externalizing behavior (β = 0.07; SD = 0.008). For the multi-PGS_cognition_, PGS for educational attainment (β = −0.052; SD = 0.011) and PGS for intelligence quotient (β = −0.08; SD = 0.027) were negatively associated with externalizing behavior. Also, PGS for suicide attempt (β = 0.02; SD = 0.009), risk tolerance (β = 0.03; SD = 0.01), smoking cessation (β = 0.04; SD = 0.007), number of sexual partners (β = −0.06; SD = 0.013), cigarettes per day (β = 0.02; SD = 0.011), lifetime cannabis use (β = −0.02; SD = 0.012) and alcohol intake frequency (β = −0.06; SD = 0.013) were all consistently contributing to externalizing behavior and improved the prediction accuracy of the multi-PGS_health-risk_behaviors_. In contrast, neither multi-PGS_personality_ nor multi-PGS_non-mental_diseases_ improved the prediction accuracy over the single-PGS model and did not integrate any consistent single PGS contributing to externalizing behavior.

The permutation test revealed that multi-PGS_health-risk_behaviors_ achieved the highest improvement in prediction accuracy compared to the other multi-PGSs, with its R^2^_adj_ being 18.75 SD away from the mean of its null distribution (**Supplementary Figure 2**). Multi-PGS_cognition_ and multi-PGS_mental-health_ also showed notable improvement, with R^2^_adj_ values of 15.96 and 15.04 SD from their respective distribution means, respectively (**Supplementary Figure 2**), while multi-PGS_environment_ contributed meaningfully to the prediction of externalizing behavior but its improvement was slightly lower, with an R^2^_adj_ of 9.47 SD away from the mean of its distribution (**Supplementary Figure 2**). Conversely, multi-PGS_personality_ and multi-PGS_non-mental_diseases_ showed minimal contribution to the prediction of externalizing behavior, with R^2^_adj_ values of less than or around 1 SD away from the mean of their distributions (**Supplementary Figure 2**).

### Moderation effect by SES

SES was negatively associated (β = −0.011; p-value = 9.33 x 10^-15^) with externalizing behavior and interacted negatively with multi-PGS_environment_ (p-value: 5.28 x 10^-4^), multi-PGS_mental-health_ (p-value: 1.27 x 10^-2^), multi-PGS_cognition_ (p-value: 7.9 x 10^-4^) and multi-PGS_health-risk_behaviors_ (p-value: 2.56 x 10^-3^) in their relationship with externalizing behavior (**Figure 3, Supplementary Figure 3**). This means that the lower the individual’s SES, the stronger the influence of these four multi-PGSs on externalizing behavior.

**Figure 3.**
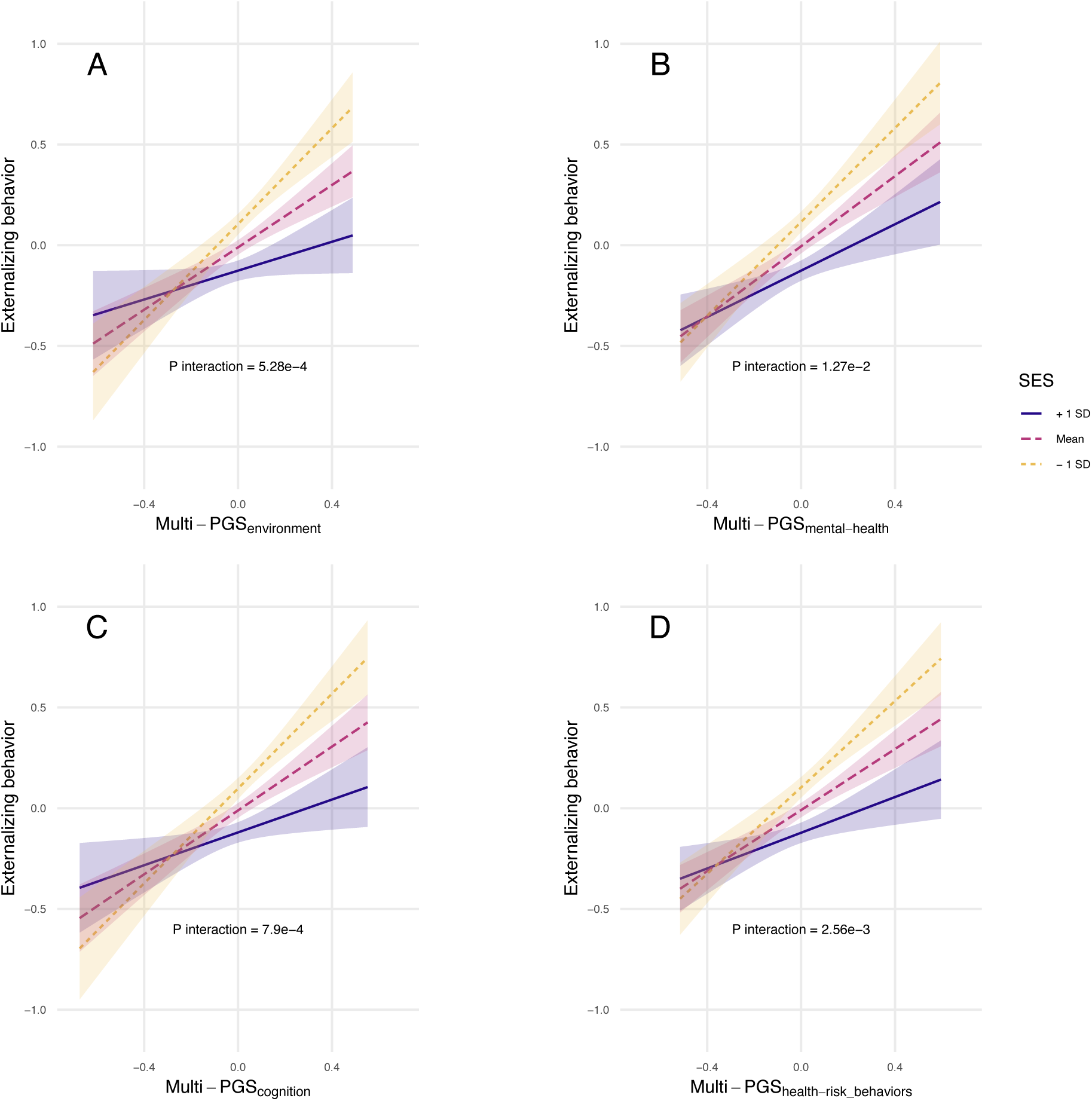
Interaction between SES and the multi-PGSs model predicted values on externalizing behavior. Predicted externalizing behavior is shown across a range of multi-PGS score values for different levels of SES: mean (red, dashed line with longer dashes), mean + 1 SD (blue, solid line) and mean – 1 SD (yellow, dashed line with shorter dashes). Shaded areas indicate the 95% confidence intervals for predicted externalizing behavior. Positive slope indicates that increased multi-PGSs are associated with higher scores for externalizing behavior. Interaction P-value corresponds to the results obtained in the meta-analysis considering the five partitions.

## DISCUSION

Through a hypothesis-driven, categorized multi-PGS framework we incorporated PGSs across four categories (environment, mental-health, cognition and health-risk behaviors) to PGS_externalizing_ and substantially improved the phenotypic variance explained of externalizing behavior among schoolchildren in 20.9%, 37.6%, 35% and 34.2%, respectively, over the single-PGS model based solely on PGS_externalizing_. Furthermore, an interaction between these multi-PGS and SES was found indicating that the genetic influence on externalizing behavior was amplified at lower levels of SES. Overall, our findings support the use of multi-PGS approaches as a promising method to increase variance explained and gain insights into the etiology of the studied condition.

By combining multiple PGSs into multi-PGS models, we increased the total variance explained and improved prediction performance by adding and decomposing the genetic signal into more specific components and redistributing shared variances across multiple predictors. As expected, PGS_externalizing_ was the strongest contributor to externalizing behavior across all multi-PGS models. Moreover, in the multi-PGS models that did not improve the prediction accuracy over the single-PGS model (i.e., multi-PGS_personality_ and multi-PGS_non-mental_diseases_), PGS_externalizing_ was the only consistent contributor and its weights were similar to those obtained in the single-PGS model. Conversely, compared with the single-PGS model, the weight of PGS_externalizing_ decreased in multi-PGS_environment_, multi-PGS_mental-health_ and multi-PGS_cognition_, where additional consistent PGSs contributed to improve the prediction accuracy of externalizing behavior. These findings suggest that, beyond their genetic contribution to externalizing behavior, additional PGSs may also capture part of the variance attributed to the PGS_externalizing_ in the single-PGS model. Among consistent PGSs that contributed to increase prediction accuracy of externalizing behavior in these multi-PGS models we find disorders that are classified as externalizing (i.e., ADHD), as well as educational attainment and intelligence quotient, two cognition-related traits displaying negative genetic correlation with externalizing behavior (Karlsson Linnér et al., 2021). This finding adds further evidence supporting associations between externalizing problems in childhood and adolescence and difficulties in academic competence and future school dropout, relationships that are known to be partially explained by shared genetic effects (Vuoksimaa et al., 2021; Veldman et al., 2014; Lewis et al., 2017; Esch et al., 2014).

The genetic liability of social outcomes, specifically, job involves heavy manual or physical work and average total household income, also enhanced the polygenic prediction of externalizing behavior. These results are consistent with prior research supporting that lower household income increases emotional and behavioral problems, as well as with the negative genetic correlation between household income and externalizing behavior (Huisman et al., 2010; Karlsson Linnér et al., 2021). It aligns with our finding showing a negative association between SES and externalizing behavior and provide additional evidence that social inequalities shape children’s behavior.

In the health-risk behaviors category, the substantial improvement in the prediction accuracy of externalizing behavior over the single-PGS model was attributable to specific PGSs for risky behaviors that show an externalizing component (i.e., suicide attempt, risk tolerance and number of sexual partners), as well as behaviors related to self-regulation (i.e., cigarettes per day, smoking cessation, alcohol intake frequency and lifetime cannabis use), many of which are known to exhibit substantial genetic overlap with externalizing behavior (Ronald et al., 2021; Albiñana et al., 2023). Consistent with prior findings, these PGSs contributed to the prediction of externalizing behavior and showed effects in the expected direction, with the exception of PGS_number_of_sexual_partners_ (Fujikane et al., 2024; Karlsson Linnér et al., 2019; Baselmans et al., 2022). This finding may reflect Simpson’s paradox, a phenomenon where the effect of a predictor (PGS_number_of_sexual_partners_) reverses direction when combined with other correlated predictor (PGS_externalizing_). The negative coefficient observed for PGS_number_of_sexual_partners_ in the muti-PGS model may not indicate a reduction in risk of externalizing behavior. Rather, it may reflect shared genetic variance with PGS_externalizing_. Once PGS_externalizing_ is included in the model, the unique variance captured by PGS_number_of_sexual_partners_ may represent a distinct genetic signal (e.g., related to sociability), that exhibits an opposite direction of association with externalizing behavior.

The impact of the multi-PGSs on externalizing behavior was moderated by SES, with the polygenic contribution to externalizing problems being exacerbated among individuals with low SES. Previous studies assessing GxE, regarding externalizing behaviors, have mainly focused on ADHD. Østergaard et al. did not find significant interaction between the genetic liability to ADHD and maternal/paternal history of mental disorders, education, work status, or income upon the symptoms of ADHD (Østergaard et al., 2020). Conversely, Leffa et al., exploring GxE using PGS_ADHD_ and environmental risk score (ERS), including, for example, mother education and single parent family, found significant interaction between PGS_ADHD_ and ERS on the CBCL attention problems syndrome scale and on the SDQ hyperactivity scores (Leffa et al., 2024). Overall, as these studies highlight the complex interdependence between SES and mental health, SES emerges as an essential factor in psychiatric genetics research, where further investigation is needed to better understand the relationship between SES and health disparities.

Although our results support that combining multiple PGSs has potential to improve outcome prediction, this study should be interpreted under some limitations. First, the contribution of predictive PGSs to externalizing behavior was modest, which was expected given that current PGSs partially capture heritability and explain a limited portion of the trait variance. To address multicollinearity among predictors, we employed an L1-penalized regression model, which allowed us to select the most predictive PGSs from subsets of correlated scores within each category. Although individual effect sizes were small, including multiple contributing PGSs enhanced overall predictive accuracy and helped further disentangle the genetic architecture of externalizing behavior. Second, our categorization strategy may have influenced in the assignment of the final PGS’ weights. Several selected predictors across categories may share genetic background and might have been excluded under a broader and more comprehensive study design. Similarly, combining this hypothesis-driven categorized approach with L1-penalized regressions, may have led to the exclusion of relevant traits and disorders, particularly those related with risk-taking behaviors or derived from overlapping GWAS summary statistics (e.g., externalizing behavior, risk tolerance and number of sexual partners). These phenotypes may contribute to externalizing behavior but also share genetic architecture with other predictors already included in the multi-PGS models. Third, although we cannot rule out that some relationships may not be linear, we used a lasso model that assumes linear associations of the outcome with the PGSs and covariates following the design described in Albiñana et al. (Albiñana et al., 2023). Fourth, we examined the moderating effects of SES on externalizing behavior and provided key insights for identifying vulnerable individuals in the general population. However, the relatively small sample size of the cohort limited our ability to investigate whether other factors, such as sex or age, may also interact with the relationship between the multi-PGSs and externalizing behavior. Finally, participants were exclusively individuals of European descent. Given that allele frequencies, linkage disequilibrium patterns, and effect sizes of common polymorphisms vary with ancestries (Martin et al., 2017), genetic findings based on common variant may not translate well across populations (Martin et al., 2019; Mills & Rahal, 2019).

## CONCLUSION

In conclusion, this study provides new insights into the genetic architecture of externalizing behavior in a population-based cohort of children and adolescents and further supports the utility of incorporating multiple PGSs into predictive models to improve accuracy, increase the proportion of variance explained, and maximize the predictive utility of PGSs. Additionally, by assessing GxE, we contribute to understanding the complex relationship between SES and mental health outcomes, highlighting its impact on youth.

## Data Availability

Raw data from this article are not publicly available because of limitations in ethical approvals, and the summary data will be available from the corresponding author upon reasonable request.

## ACKNOWLEDGMENTS

The authors are grateful to the families, students, and staff of the public primary schools (i.e., Joan Maragall, Maria Borés, Marquès de la Pobla, Martinet, Pins del Vallès, Puiggraciós, Sant Jordi, Ramon Llull, Rivo Rubeo, Tagamanent, and Teresa Berguedà), public secondary schools (i.e., Angeleta Ferrer i Sensat, Antoni Pous i Argila, Cal Gravat, Duc de Montblanc, Institut del Ter, Jaume Callís, Lacetània, Lluís de Peguera, Molí de la Vila, Montsuar, Pius Font i Quer, Vallbona d’Anoia, and Vil·la Romana), and private schools (i.e., Airina, L’Ave Maria, Casals–Gràcia, Episcopal Lleida, La Farga, FEDAC Manresa, FEDAC Vic, Garbí Pere Vergés Esplugues, Institució Igualada, Joviat, Oms i de Prat, Pies Mataró, Pureza de María, Regina Carmeli, Sagrats Cors Centelles, La Salle Manlleu, La Salle Manresa, Sant Miquel dels Sants, Thau Barcelona, and Vedruna Escorial Vic) who kindly contribute to this research. The genotyping service was carried out at the Genotyping Unit-CEGEN in the Spanish National Cancer Research Centre (CNIO), supported by Instituto de Salud Carlos III (ISCIII), Ministerio de Ciencia e Innovación. CEGEN is part of the initiative IMPaCTGENóMICA (IMP/00009) cofunded by ISCIII and the European Regional Development Fund (ERDF).

