## Supplementary Figures for "Multi-polygenic scores for externalizing behavior in schoolchildren"

**SUPPLEMENTARY MATERIAL**


1. Supplementary Figure 1. PGS correlation plots……………………………………………………………………….1
2. Supplementary Figure 2. $R^2_{adj}$ null distribution and multi-PGSs' observed $R^2_{adj}$……………………….5
3. Supplementary Figure 3. Forest plots showing interaction effects between SES and multi-PGSs on externalizing behavior……………………………………………………………………………………………7

A)

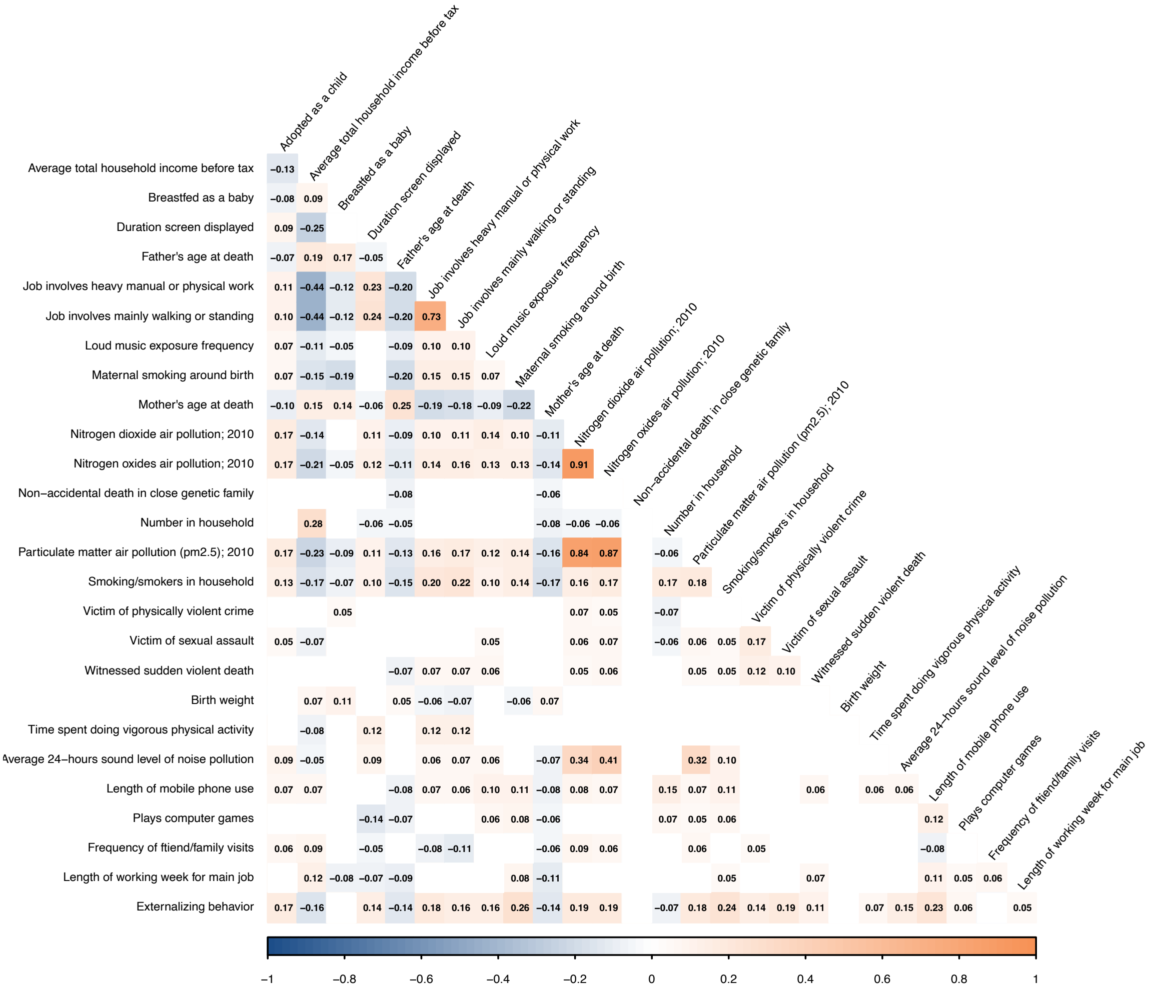
Adopted as a child
Average total household income before tax
Breastfed as a baby
Duration screen displayed
Father's age at death
Job involves heavy manual or physical work
Job involves mainly walking or standing
Loud music exposure frequency
Maternal smoking around birth
Mother's age at death
Nitrogen dioxide air pollution; 2010
Nitrogen oxides air pollution; 2010
Non−accidental death in close genetic family
Number in household
Particulate matter air pollution (pm2.5); 2010
Smoking/smokers in household
Victim of physically violent crime
Victim of sexual assault
Witnessed sudden violent death
Birth weight
Time spent doing vigorous physical activity
Average 24−hours sound level of noise pollution
Length of mobile phone use
Plays computer games
Frequency of ftiend/family visits
Length of working week for main job
Externalizing behavior
−1
−0.8
−0.6
−0.4
−0.2
0
0.2
0.4
0.6
0.8
1

B)

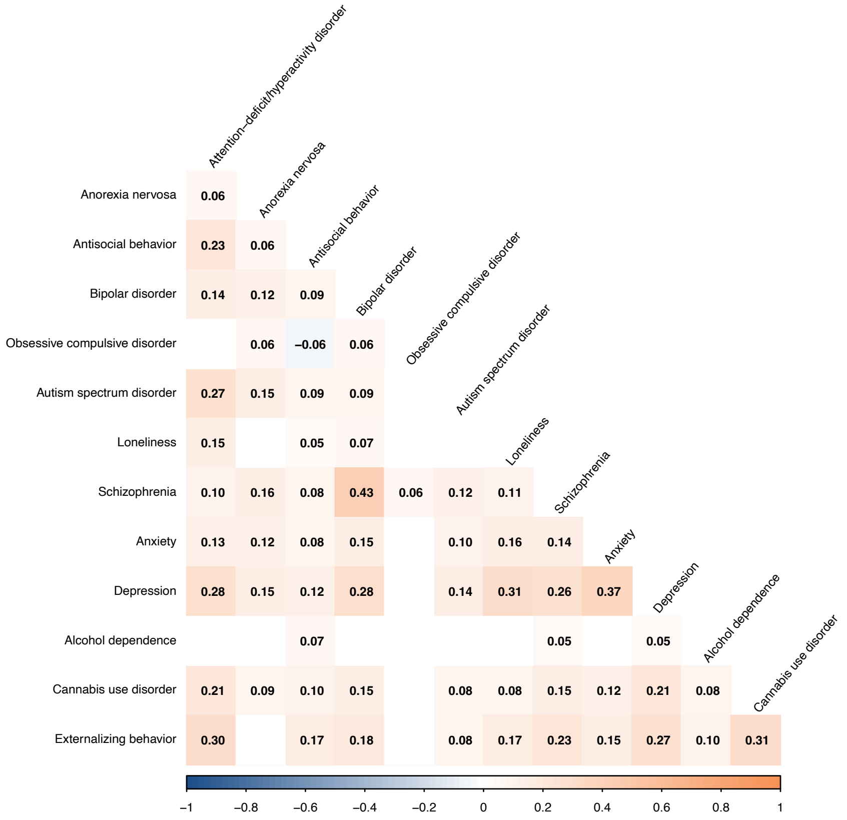
Attention−deficit/hyperactivity disorder
Anorexia nervosa
Antisocial behavior
Bipolar disorder
Obsessive compulsive disorder
Autism spectrum disorder
Loneliness
Schizophrenia
Anxiety
Depression
Alcohol dependence
Cannabis use disorder
Externalizing behavior
−1
−0.8
−0.6
−0.4
−0.2
0
0.2
0.4
0.6
0.8
1

C)

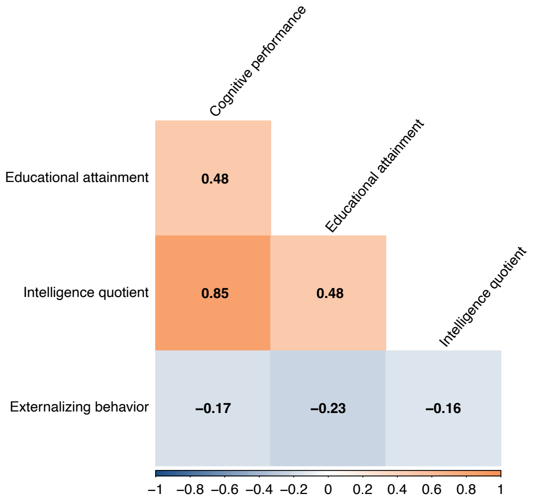
Cognitive performance
Educational attainment
Intelligence quotient
Educational attainment
Intelligence quotient
Externalizing behavior
0.48
0.85
0.48
−0.17
−0.23
−0.16
−1 −0.8 −0.6 −0.4 −0.2 0 0.2 0.4 0.6 0.8 1

D)

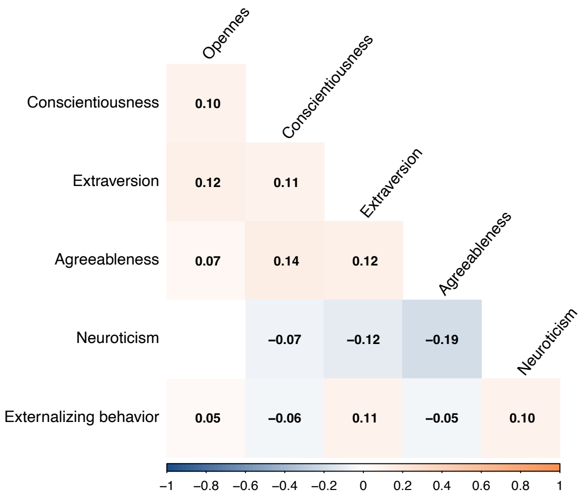
Opennes
Conscientiousness
Extraversion
Agreeableness
Neuroticism
Conscientiousness
Extraversion
Agreeableness
Neuroticism
Externalizing behavior
0.10
0.12
0.11
0.07
0.14
0.12
−0.07
−0.12
−0.19
0.05
−0.06
0.11
−0.05
0.10
−1 −0.8 −0.6 −0.4 −0.2 0 0.2 0.4 0.6 0.8 1

E)

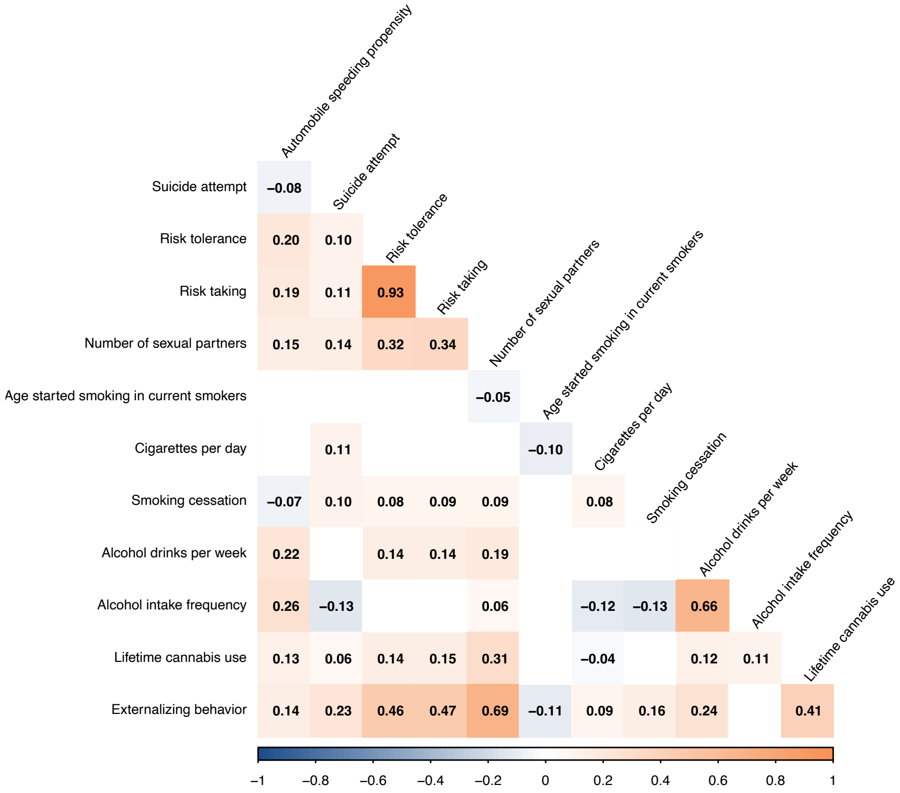
Automobile speeding propensity
Suicide attempt
Risk tolerance
Risk taking
Number of sexual partners
Age started smoking in current smokers
Cigarettes per day
Smoking cessation
Alcohol drinks per week
Alcohol intake frequency
Lifetime cannabis use
Suicide attempt
Risk tolerance
Risk taking
Number of sexual partners
Age started smoking in current smokers
Cigarettes per day
Smoking cessation
Alcohol drinks per week
Alcohol intake frequency
Lifetime cannabis use
Externalizing behavior
−0.08
0.20 0.10
0.19 0.11 0.93
0.15 0.14 0.32 0.34
−0.05
0.11 −0.10
−0.07 0.10 0.08 0.09 0.09 0.08
0.22 0.14 0.14 0.19
0.26 −0.13 0.06 −0.12 −0.13 0.66
0.13 0.06 0.14 0.15 0.31 −0.04 0.12 0.11
0.14 0.23 0.46 0.47 0.69 −0.11 0.09 0.16 0.24 0.41
−1 −0.8 −0.6 −0.4 −0.2 0 0.2 0.4 0.6 0.8 1

F)

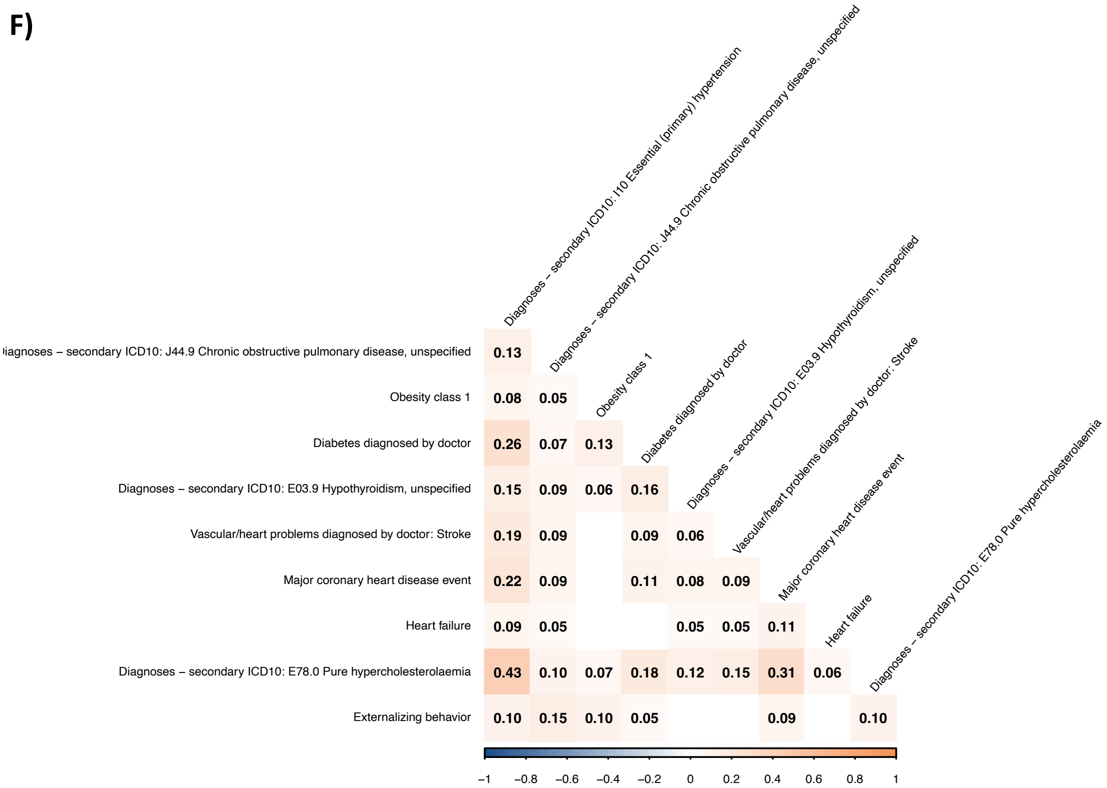

Diagnoses – secondary ICD10: I10 Essential (primary) hypertension
Diagnoses – secondary ICD10: J44.9 Chronic obstructive pulmonary disease, unspecified
Obesity class 1
Diabetes diagnosed by doctor
Diagnoses – secondary ICD10: E03.9 Hypothyroidism, unspecified
Vascular/heart problems diagnosed by doctor: Stroke
Major coronary heart disease event
Heart failure
Diagnoses – secondary ICD10: E78.0 Pure hypercholesterolaemia
Externalizing behavior
−1 −0.8 −0.6 −0.4 −0.2 0 0.2 0.4 0.6 0.8 1


G)

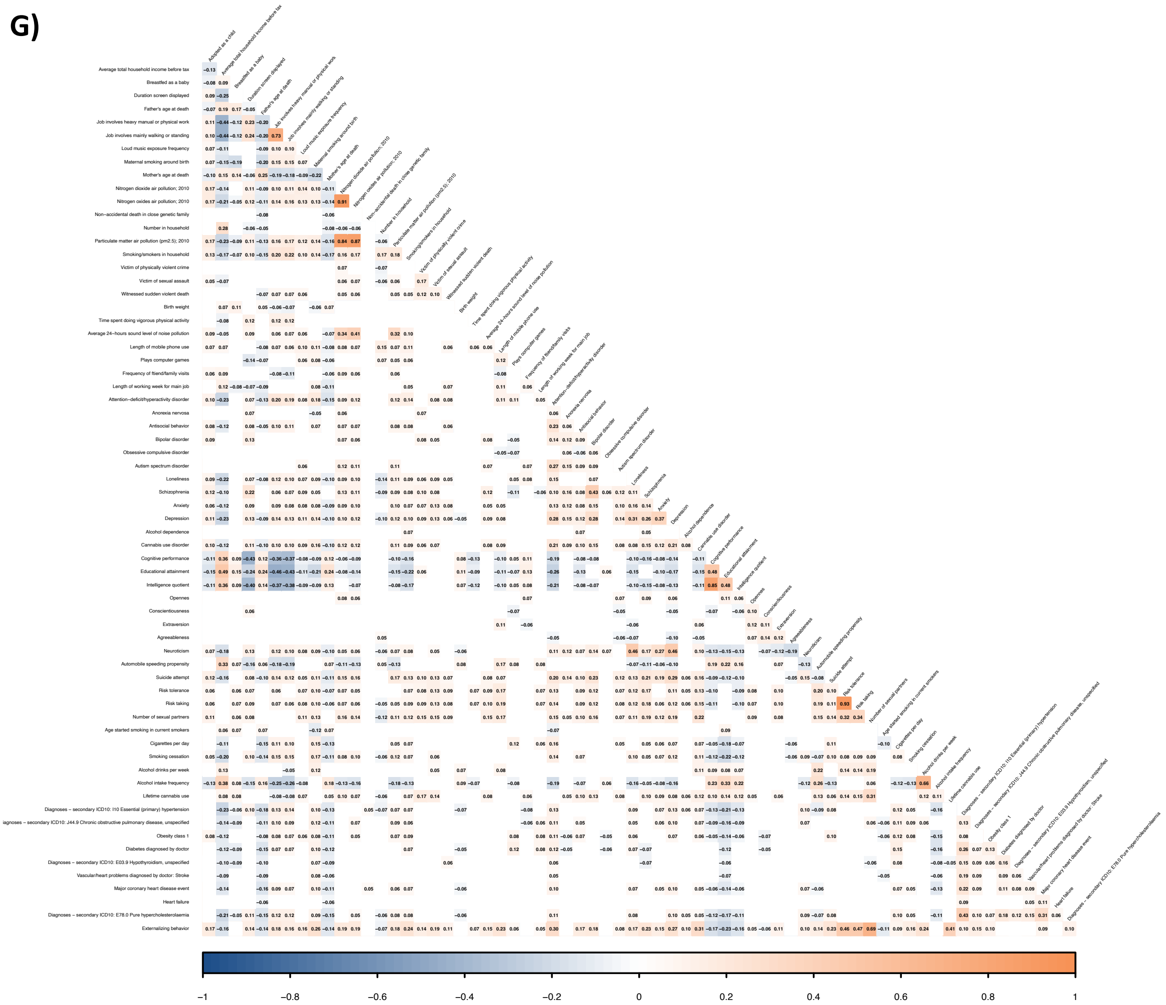

**Supplementary Figure 1. PGS correlation plots.** Pearson's correlation between PGSs of the **A)** environmental **B)** mental-health **C)** cognition **D)** personality **E)** health-risk behaviors **F)** non-mental health category. **G)** Pearson's correlation between all PGSs used. Non-significant (Bonferroni) correlations are in blank, positive correlations are shown in orange and negatives in blue.

A)

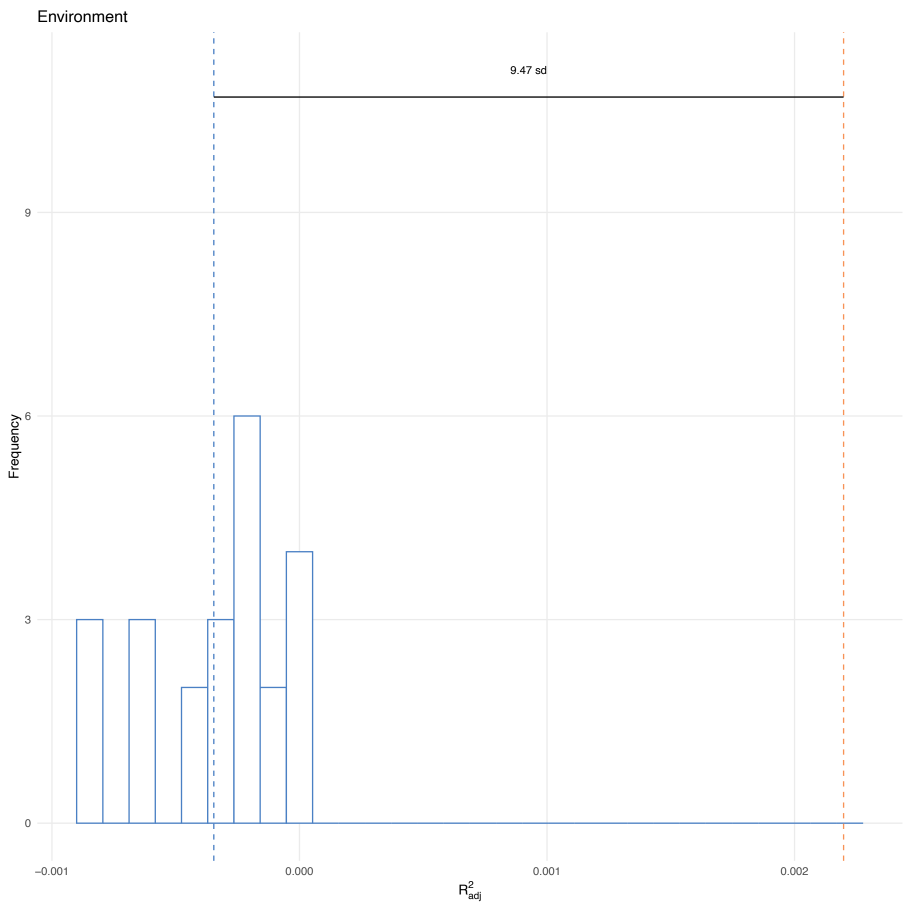
Environment
9.47 sd
Frequency
9
6
3
0
−0.001
0.000
0.001
0.002
$R^2_{adj}$

B)

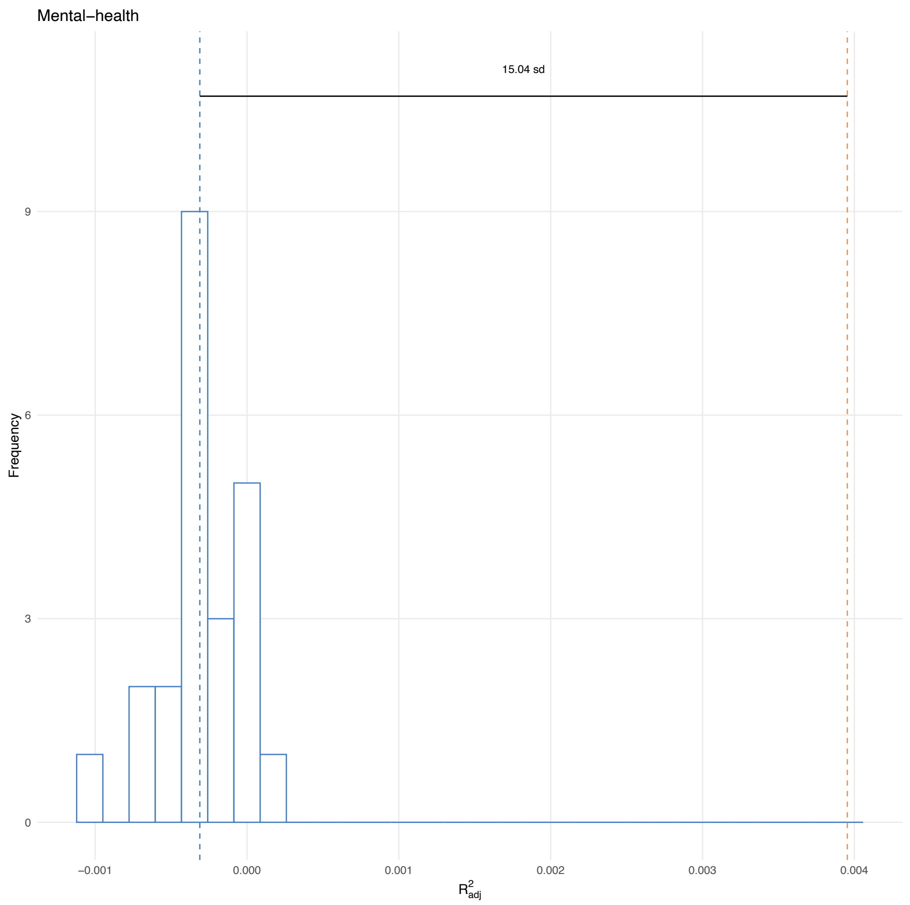
Mental−health
15.04 sd
Frequency
9
6
3
0
−0.001
0.000
0.001
0.002
0.003
0.004
$R^2_{adj}$

C)

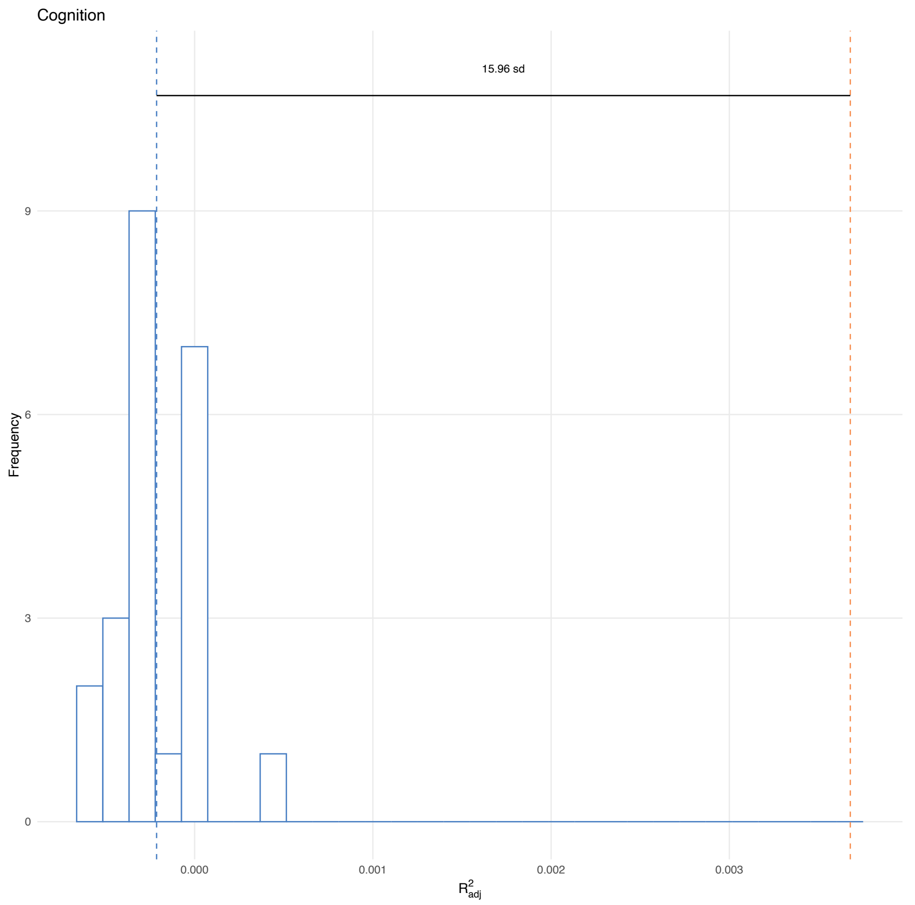
Cognition
15.96 sd
9
6
3
0
Frequency
0.000
0.001
0.002
0.003
$R^2_{adj}$

D)

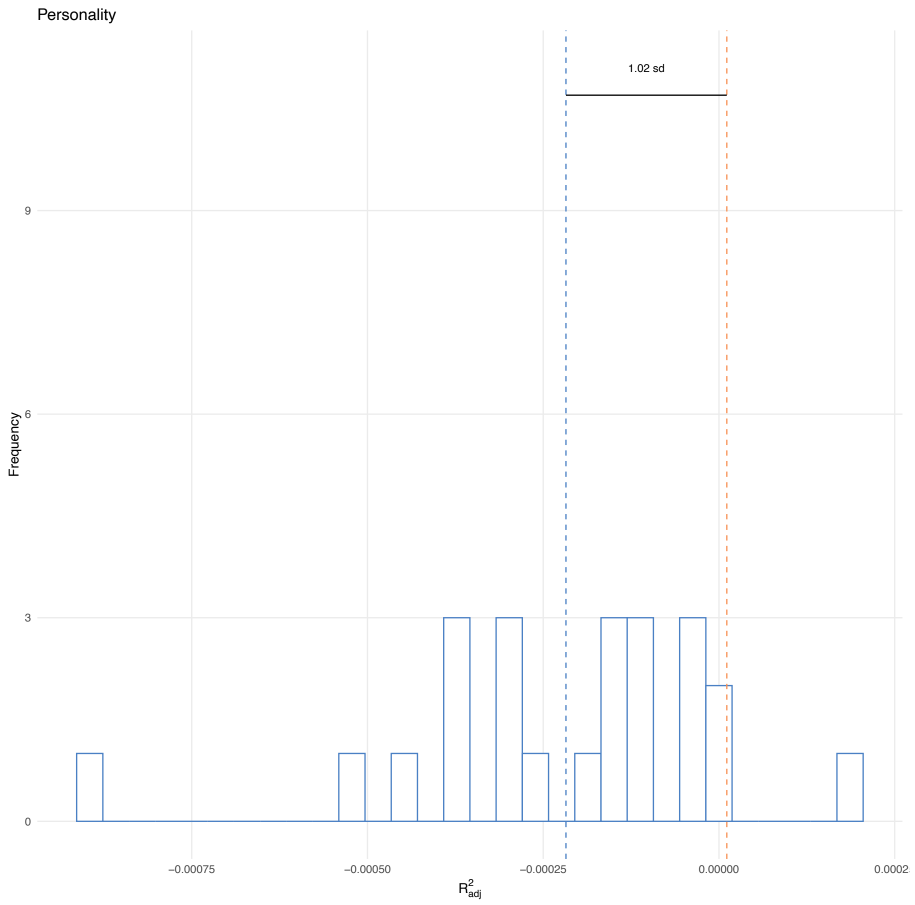
Personality
1.02 sd
9
6
3
0
Frequency
−0.00075
−0.00050
−0.00025
0.00000
0.0002
$R^2_{adj}$

**E)**

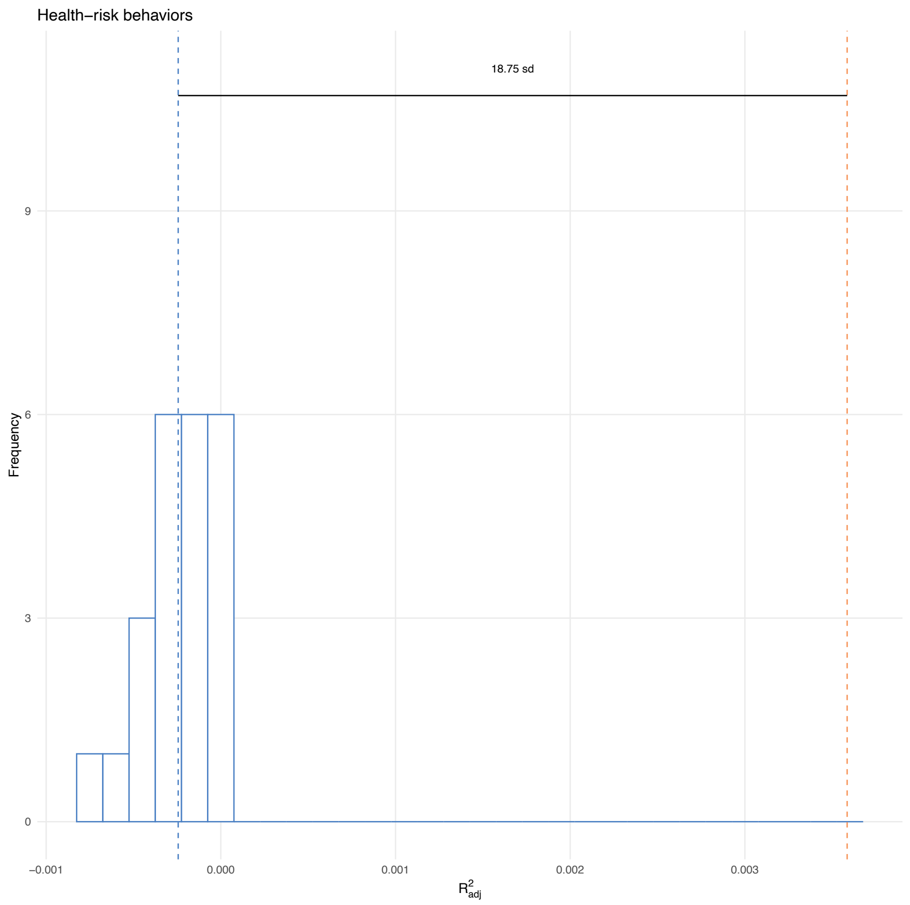


**F)**

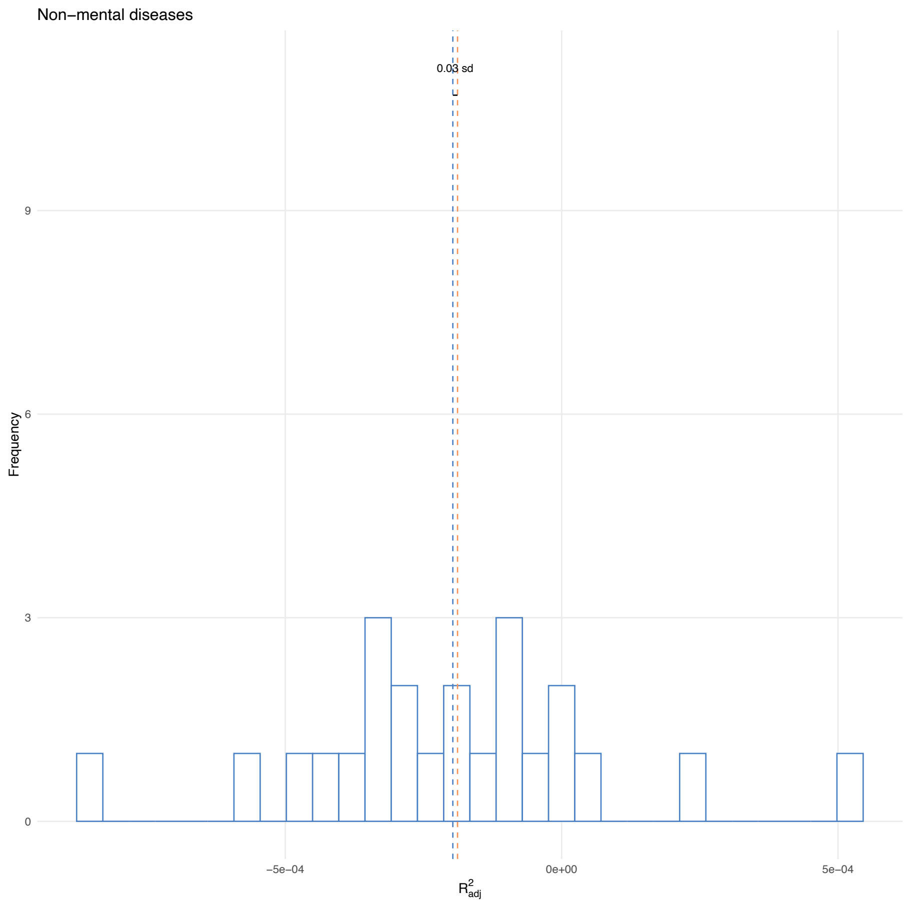


**Supplementary Figure 2. $R^2_{adj}$ null distribution and multi-PGSs' observed $R^2_{adj}$.** $R^2_{adj}$ null distribution obtained from the permutation test of **A)** multi-$PGS_{environment}$ **B)** multi-$PGS_{mental\text{-}health}$ **C)** multi-$PGS_{cognition}$ **D)** multi-$PGS_{personality}$ **E)** multi-$PGS_{health\text{-}risk\_beahviors}$ **F)** multi-$PGS_{non\text{-}mental\_diseases}$ is shown in blue. The dashed blue line indicates the mean of the null distribution and the dashed orange line the observed $R^2_{adj}$. The number of null distribution's standard deviations (SD) between the observed $R^2_{adj}$ and the mean of the null distribution are also presented.

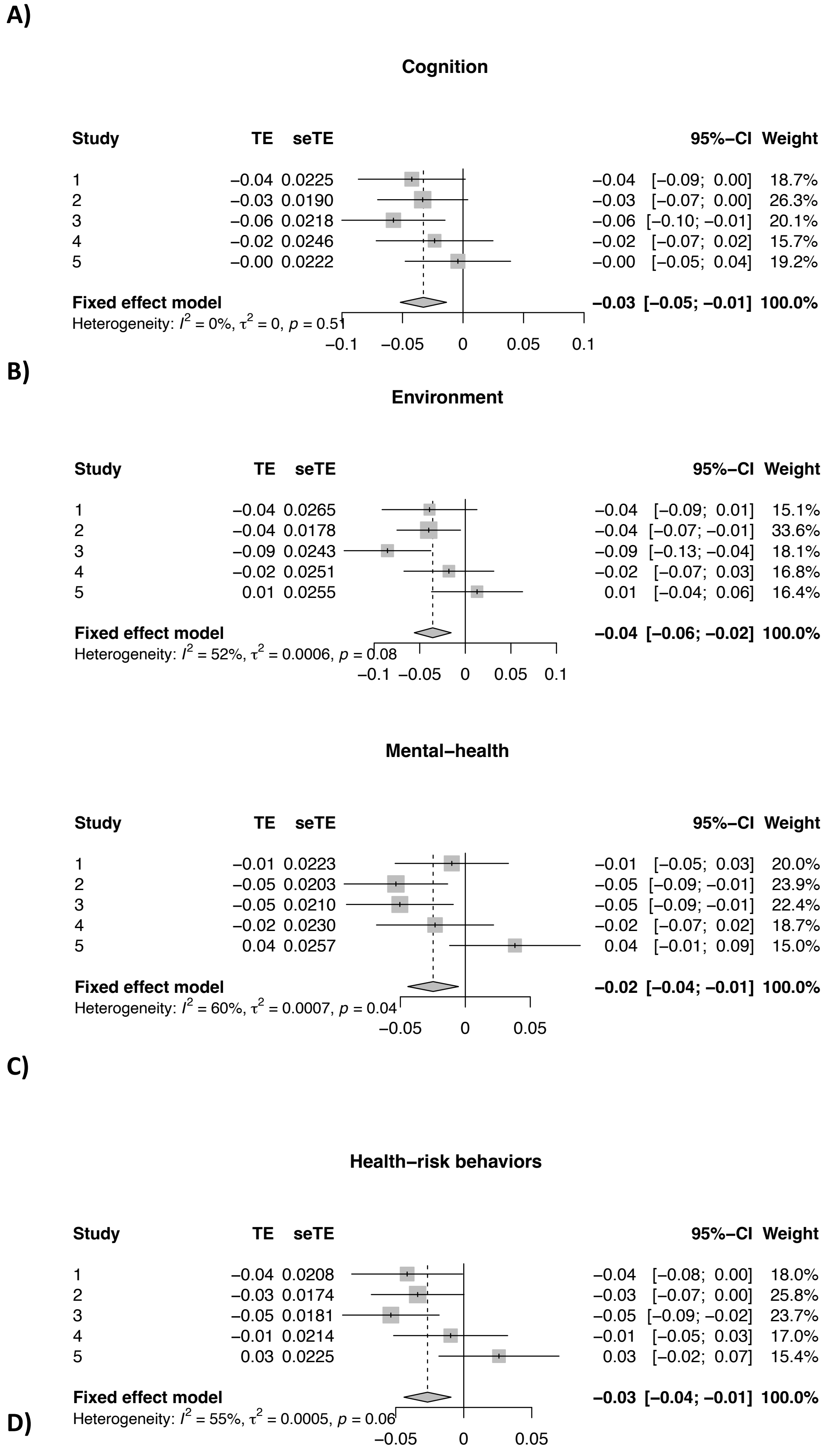


**Supplementary Figure 3. Forest plots showing interaction effects between SES and multi-PGSs on externalizing behavior.** Estimated interaction effect between SES and the five partitions of **A)** multi-$PGS_{environment}$ **B)** multi-$PGS_{mental\text{-}health}$ **C)** multi-$PGS_{cognition}$ **D)** multi-$PGS_{health\text{-}risk\_beahviors}$ on externalizing behavior, along with corresponding 95% confidence intervals. Negative values indicate stronger polygenic effects in individuals with lower SES. The pulled result is shown with a diamond.
